# Is an active hospital microbiology laboratory cost-effective in a resource-limited setting? - a case study from Timor-Leste

**DOI:** 10.1101/2024.05.14.24307355

**Authors:** Cherry Lim, Myo Maung Maung Swe, Angela Devine, Tessa Oakley, Karen Champlin, Pyae Sone Oo, Nevio Sarmento, Ismael Da Costa Barreto, Rodney C Givney, Jennifer Yan, Joshua R Francis, Ben S Cooper

## Abstract

Maintaining an active hospital microbiology laboratory allows definitive antibiotic treatment for bacterial infections to be given in a timely manner. This would be expected to improve patient outcomes and shorten length of hospital stay. However, many hospitals in low- and middle-income countries lack access to microbiology services, and the cost-effectiveness of an active microbiology service is unknown. We constructed a decision tree and performed a cost-effectiveness model analysis to determine whether maintaining an active microbiology laboratory service would be cost-effective in Timor-Leste, a lower middle-income country. The analysis was informed by local microbiology data, local patient treatment costs, results of an expert elicitation exercise and data from literature reviews. The results indicate that there is a high probability that maintaining an active microbiology laboratory is a cost-effective intervention that would both improve patient outcomes and reduce net costs (due to reduced intensive care admissions and potential costs of resistant infections) compared to no microbiological testing, especially for the hospitalised paediatric patients with suspected primary bacteraemia. This remained true under various one-way sensitivity analyses, including when accuracy of microbiological testing is low, prevalence of bacterial infection among patients with suspected bloodstream infection was high, and prevalence of antibiotic resistance was high.

## Introduction

The emergence of antimicrobial resistance (AMR) is a global health threat, and the burden of AMR is highest in low-and middle-income countries (LMICs) with limited healthcare resources to respond to and monitor the spread of resistance.^1^ Timor-Leste is a relatively new country, gaining formal independence from Indonesia in 2002 with a current population of 1.3 million, predominantly rural (74%).^2^ Hospital Nasional Guido Valadares (HNGV) is the tertiary referral hospital for the country, located in the capital city of Dili with a 340-bed capacity.^4^ HNGV receives a microbiology service from the adjacent referral laboratory, Laboratorio Nacional de Saude. Previous studies have indicated a high rate of AMR in Timor-Leste.^5–7^

A previous analysis of surveillance by blood culture in low to middle income countries in Africa to inform empirical antibiotic therapy policy in sepsis concluded that this intervention was cost-effective for this purpose, but did not consider whether routine use of blood cultures for microbiologically-informed targeted antibiotic treatment in the patients tested was also economically rational. ^8^

The aim of the current study was to determine if, in Timor-Leste, a low-middle income country, blood culture and sensitivity testing to guide antibiotic therapy for the individual patient with sepsis at the HNGV tertiary referral hospital is cost-effective.

## Methods

While local hospital active microbiological testing may provide many benefits, including providing information to support infection prevention and control programmes and to develop local empirical antibiotic guidelines, we consider only the health benefits arising from improved treatment of suspected bloodstream infection in hospital inpatients in this analysis. In this model, we assume the patient is hospitalised and is suspected to have a bloodstream infection.

### Decision tree model

We consider two comparison arms and calculate expected costs and health-related outcomes including mortality and length of hospitalisation associated with each arm (Figure 1). The decision trees were drawn in SilverDecisions 1.2.0 (https://silverdecisions.pl/), and all analyses were performed using R (version 4.0.5; The R Foundation for Statistical Computing, Vienna, Austria).

**Figure 1.**
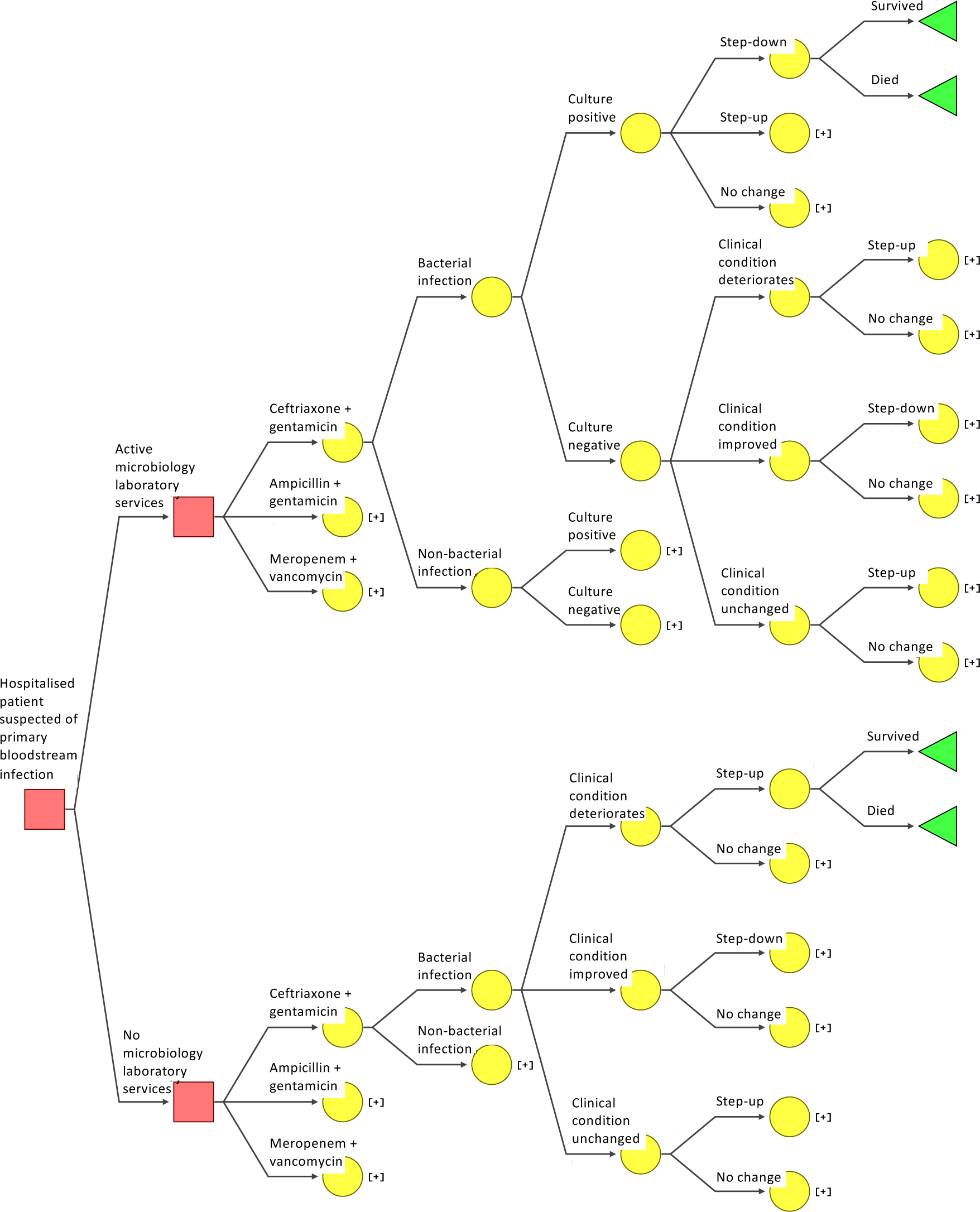
Decision tree to estimate the cost-effectiveness of an active microbiology laboratory surveillance system compared to no microbiological testing. Squares indicate decision nodes, circles indicate chance nodes, and triangles indicate the end point. Collapsed branches of the tree are indicated by “[+]” and represent repeats of those shown at corresponding nodes on other branches of the tree.

In the active microbiology laboratory testing system arm, a patient with suspected bloodstream infection will have one set of blood cultures (two bottles) taken, and be started on empirical antibiotic therapy before any potentially causative organism is identified. If bacterial pathogens are identified from those cultures, then definitive therapy based on the antibiotic susceptibility of the organisms may lead to the antibiotic regimen being changed. In the second arm, in which there is no microbiological testing, the patient would again be empirically treated, but any changes in treatment would be driven by clinical condition alone, without microbiological testing results. In both comparison arms, we consider three antibiotic regimens recommended at the time of writing in local guidelines for empirical antibiotic use in Timor-Leste: ceftriaxone and gentamicin, ampicillin and gentamicin, and meropenem and vancomycin.^9^ The first two antibiotic recommendations are also consistent with WHO recommendations for empiric antibiotics in clinical sepsis of unknown origin in adults and children respectively.^10^ Meropenem and vancomycin is a common antibiotic combination used in Timor-Leste for the critically unwell patient.

The observed microbiology testing result may not always represent the true infection status because the sensitivity and specificity of microbiology testing is not 100%, therefore the true infection status of a patient is assumed to be imperfectly observed.

In the active microbiology laboratory testing system arm, the microbiology testing results are assumed to depend on the true infection status of the patient and the performance characteristics of the testing procedures at the local microbiology laboratory.

We then assumed a patient underwent antibiotic treatment adjustment after three days of empirical treatment. The adjustment depends on which comparison arm they are under (Figure 1). For patients in the active microbiology laboratory arm, in which blood culture was performed, antibiotic treatment changes would depend on the results of blood culture. For patients in the no microbiology testing service arm, antibiotic treatment changes would depend on changes in clinical condition. We consider three possible changes to the antibiotic treatment received following initial empirical treatment:

- *stepping up*, which we define as switching to a broader spectrum or last-resort antibiotic (or antibiotic combination) based on AWaRe category, which classifies antibiotics based on their spectrum of activity and potential for resistance.^10^ An example of *stepping up* would be changing from ampicillin and gentamicin, or ceftriaxone and gentamicin, to meropenem and vancomycin.
- *stepping down*, which we define as switching to a narrower spectrum antibiotic (or antibiotic combination) or to an oral antibiotic. Examples of *stepping down* include changing from meropenem and vancomycin to ceftriaxone and gentamicin; or changing from ceftriaxone and gentamicin, to an oral augmentin.
- *no change*, which we define as patients continuing to receive the same antibiotic treatment.

### Populating the model

#### Transition probabilities

Each internal node of the decision tree has probabilities associated with each of the two or more branches splitting off from the main branch. These probabilities (which, for each node, must sum to one) represent either the probabilities of different decisions being made (e.g. step up, step down, no change) or probabilities relating to the patient’s condition (e.g. clinical condition improves, deteriorates, or remains unchanged). These probabilities were derived from literature review^11–14^, analysis of local hospital microbiology data from Timor-Leste and an expert elicitation exercise performed with physicians based at HNGV in Timor-Leste. Details of the expert elicitation are in Appendix 1, and detailed input values and references are in Appendix 2. Under the active microbiology laboratory service arm, there were four further transition events. These related to i) true bacterial infection state (which is imperfectly observed and is a “latent” variable); ii) microbiological testing result; iii) change in antibiotic treatment (assumed to be influenced by the microbiological testing); and iv) treatment outcome. Under the same arm when microbiological testing gave negative culture results, changes in antibiotic treatment were assumed to be based only on the clinical condition of the patient. In the no microbiological testing scenario, changes in antibiotic treatment were assumed to always depend only on the clinical condition of the patient regardless of the type of empirical antibiotic treatment received.

#### Costs

Model input values for costs came from national procurement drug costing data. Costs for microbiology testing were derived from local data on laboratory equipment costs, antimicrobial-susceptibility testing (AST) costs, and maintenance costs of an active microbiology laboratory. The costs of healthcare for patients were extracted from literature review and local staffing costs. Probability distributions were constructed to account for the uncertainty in costs of healthcare for patients. The maintenance costs of an active microbiological testing laboratory were derived from local data provided by the local study investigator team. Appendix 2 shows the detailed cost breakdown and sources of data. All costs were in 2022 US dollars ($). In brief, the per sample cost of maintaining an active microbiology laboratory could be as high as $34.53 (used in the model for the baseline scenario to ensure the results reflect a conservative scenario where costs of maintaining an active microbiology laboratory are high) without external funding support, and as low as $24.96 (used in the sensitivity analysis to reflect a scenario with some external funding support). The cost of one positive blood culture bottle (including microbiology culture, identification, AST, and other laboratory costs) was $31.90 and that of one negative blood culture bottle was $5.80. We assume that each patient with suspected primary bloodstream infection would have two blood culture bottles taken for microbiological testing in the active microbiology laboratory service arm. The treatment cost of ceftriaxone and gentamicin, ampicillin and gentamicin, and meropenem and vancomycin per day was $1.76, $3.04, and $35.16, respectively. The cost of clinical deterioration was calculated based on the cost of patient care in an intensive care unit (ICU), and that of clinical improvement was based on the cost of patient care in a general medicine ward. These costs were estimated from literature reviews with local hospital resourcing data (Appendix 2). We also included the cumulative costs of antimicrobial resistance per full course of antibiotic treatment based on estimates from a previous study.^15^

#### Measurement of patient outcome

Disability-adjusted life-years (DALYs) were calculated from mortality, length of hospitalisation, disability weights, and number of years lost for deaths. DALYs were calculated by estimating the sum of the years of life lost due to premature mortality (YLLs) and years of life lost due to time lived in states of less than full health, which is also known as the years of healthy life lost due to disability (YLDs). In other words, one DALY was the loss of one year of full health. The YLLs were calculated as the number of deaths multiplied by a loss function specifying the years lost for deaths as a function of the age at which death occurs, which is 11 years for patients in the age range of 55-59 years given the life expectancy of Timor-Leste population at birth of 70 years.^12^ This age group was chosen because this is one of the most common age groups for suspected bloodstream infections in Timor-Leste (unpublished data from National Laboratory Timor-Leste). YLDs were calculated from the length of hospitalisation after day of diagnosis with suspected bloodstream infection multiplied by a disability weight. The disability weight of severe acute episode of infectious disease used was 0.133 for adults based on the Global Burden of Disease (GBD) Study estimates.^13^ Patient mortality and length of hospitalisation given a specific trajectory of events were estimated based on evidence and inputs from experts working in hospitals in Timor-Leste. Details of outcomes, sources of data, and justification for the values are in Appendix 3. In addition, we performed a separate analysis with the same model but restricted to patients less than four years old, as prevalence of sepsis is high in this age group in the study hospital, assuming a loss function specifying the years lost for deaths as a function of the age at which death occurs was 66, and disability weight was 0.402 for patients less than 4 years old based on GBD estimates.

#### Measurement of cost-effectiveness

Results were expressed as an incremental cost-effectiveness ratio (ICER) for a hypothetical cohort of 1,000 hospitalised patients with suspected bloodstream infection. This was calculated by dividing the difference in calculated cost between the two arms by the DALYs averted through having an active microbiology laboratory service:

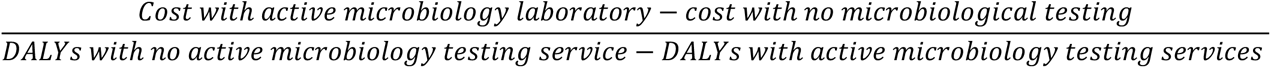

which we report as the additional cost for every DALY averted per 1,000 patients with suspected bloodstream infections.

### Interventions and scenarios considered

#### Baseline scenario

In the baseline scenario, the model assumed that i) the prevalence of true bacterial infection among suspected primary bloodstream infections was 40%; ii) the probability of pathogen detection and false positive rate of microbiological testing were 80% and 5%, respectively; and iii) the chances of having clinical condition deteriorate, improve or stay the same under different empirical treatments, and the probability of changing antibiotic prescription were derived based on the results from the expert elicitation exercise (Appendices 2 and 3). The model assumed that the sensitivity of microbiological testing reflects both how well the microbiological testing can detect the causative organism and the bacterial load. For instance, if the bacterial load in a clinical sample was high the microbiological testing would be more likely to give a positive culture result and this would then impact the outcome of the patient.

#### Deterministic sensitivity analysis

We performed stepwise deterministic sensitivity analyses to check the robustness of the conclusions under various scenarios, including: i) low and high prevalence of bacterial infection among suspected bloodstream infections; ii) high cost of maintaining microbiology laboratory per tested sample; iii) different probabilities of changing antibiotic treatments; iv) low and high risks of ICU admission; v) low and high mortalities associated with different clinical conditions; and vi) different associated costs of care for patients whose clinical conditions deteriorated post empirical antibiotic therapy. Additionally, we performed a deterministic sensitivity analysis assuming the years lost for deaths was 20 years for patients in the age range of 55-59 years based on the WHO life table.^13^ Sensitivity analysis assuming two sets of blood cultures (four bottles) was also performed.

#### Probabilistic sensitivity analysis

A probabilistic sensitivity analysis was performed using 1000 iterations.^16^ In this analysis each parameter in the decision tree was defined by an appropriate probability distribution. Specifically, the probabilities attached to each internal node that has more than two probabilities (i.e. changes in clinical conditions) were sampled from Dirichlet distributions and binomial transition probabilities were sampled from beta distributions. For patient care costs, a lognormal distribution was used to avoid negative values. Details of the distributions used are in Appendices 2 and 3.

## Results

### Baseline scenario analysis

Active microbiology surveillance reduced overall patient care costs by $161,991 (IQR: $134,583-196,461) for every 1,000 hospitalised adult patients with suspected bloodstream infection, primarily due to reduced ICU admission risk. Maintaining an active microbiology laboratory was consistently cost-saving compared to no active microbiological testing. It was most cost-saving in a setting where ampicillin and gentamicin was predominantly used as empiric treatment (Table 1). In settings where ampicillin and gentamicin, ceftriaxone and gentamicin, and meropenem and vancomycin were used as empirical antibiotic treatment, the estimated costs that an active microbiology laboratory would have saved were $277,879.00, $149,340.90, and $76,596.99 per 1,000 hospitalised patients with suspected bloodstream infections, respectively.

**Table 1.** Estimated incremental costs and disability-adjusted life years (DALYs) averted per 1,000 hospitalised adult patients from maintaining a microbiology laboratory compared to no microbiological testing under the baseline scenario. Costs are in 2022 US dollars.

| Empirical antibiotic therapy | An active laboratory | | No active microbiological testing | | Incremental costs (\$) | DALY averted | ICER |
| --- | --- | --- | --- | --- | --- | --- | --- |
| | Costs per 1,000 patients (\$) | DALY per 1,000 patients | Costs per 1,000 patients (\$) | DALY per 1,000 patients | | | |
| Ceftriaxone + gentamicin | 523,302 | 2,636 | 672,643.00 | 4,103 | -149,340.90 | 1,468 | Active laboratory dominates |
| Ampicillin + gentamicin | 537,708 | 2,640 | 815,587.20 | 4,030 | -277,879.00 | 1,390 | Active laboratory dominates |
| Meropenem + vancomycin | 717,670 | 2,479 | 794,267.20 | 4,156 | -76,596.99 | 1,677 | Active laboratory dominates |

ICER: incremental cost-effectiveness ratio

For every 1,000 hospitalised adult patients aged 50-59 years with suspected bloodstream infections, 40 (IQR: 34-48) deaths, 36 (IQR: 30-42) deaths, and 58 (IQR: 50-67) deaths would have been prevented with expanded microbiology testing capacity under the three choices of empirical antibiotic treatment (ceftriaxone and gentamicin, ampicillin and gentamicin and meropenem and vancomycin respectively) (Table 2). There is a high probability that maintaining a microbiology laboratory service would be more effective and less costly for hospitalised patient care compared to no microbiological testing in Timor-Leste (Figure 2).

**Figure 2.**
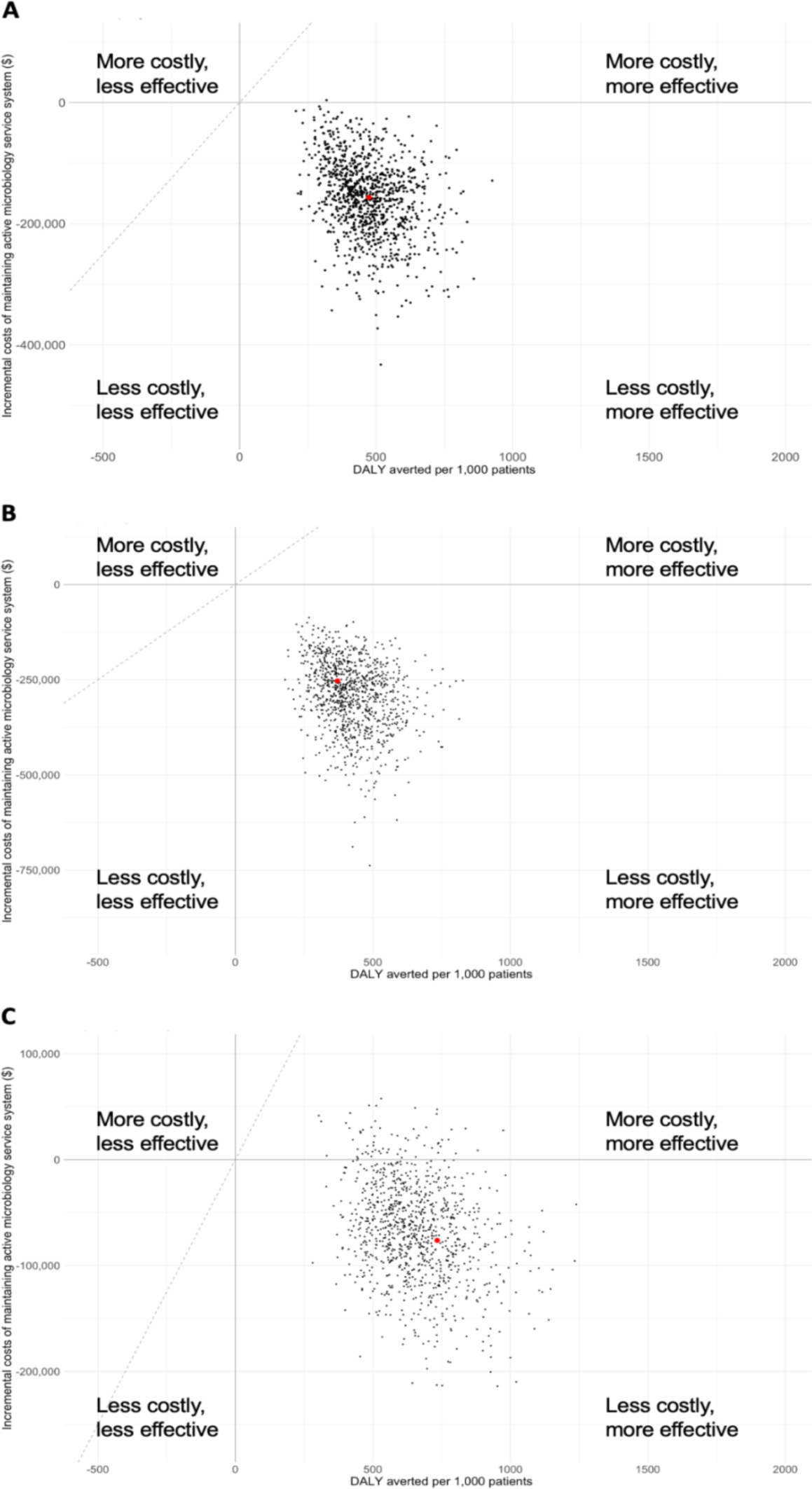
Incremental cost-effectiveness plane showing disability-adjusted life years (DALYs) averted versus incremental costs per 1,000 hospitalised patients from maintaining a microbiology laboratory compared to no microbiological testing using 1,000 model iterations when (A) ceftriaxone and gentamicin; (B) ampicillin and gentamicin; and (C) meropenem and vancomycin is used as empirical treatment. Points in black are results from the probabilistic sensitivity analysis, and the larger red point is the base case result. In the upper right quadrant, points below the broken diagonal line would be considered cost-effective for a willingness to pay to avert one DALY of $500, while points above this line would not be considered cost-effective at this threshold.

**Table 2.** Estimated costs saved and deaths averted per 1,000 hospitalised paediatric patients (under four years of age) from maintaining a microbiology laboratory compared to no microbiological testing from a probabilistic model sensitivity analysis. Costs are in 2022 US dollars.

| Empirical antibiotic | Cost saved (IQR)<br>per 1,000 hospitalised patients | Number of deaths saved (IQR)<br>per 1,000 hospitalised patients | ICER (IQR) |
| --- | --- | --- | --- |
| Ceftriaxone + gentamicin | \$152,323 (116,815-195,021) | 36 (31-43) | Active laboratory<br>dominates |
| Ampicillin + gentamicin | \$279,095 (227,477-345,003) | 33 (28-39) | Active laboratory<br>dominates |
| Meropenem + vancomycin | \$66,173 (39,090-96,921) | 50 (42-59) | Active laboratory<br>dominates |

Similar findings were observed in the population with age under 4 years (Appendix 5). For every 1,000 hospitalised patients under 4 years of age with suspected bloodstream infections, 36 (IQR: 31-43) deaths, 33 (IQR: 28-39) deaths and 50 (IQR: 42-59) deaths would have been prevented with active microbiological testing using ceftriaxone and gentamicin, ampicillin and gentamicin, and meropenem and vancomycin, respectively (Table 2). For every 1,000 hospitalised patients under 4 years of age with suspected bloodstream infections, 2,593 (IQR: 2,187-3,045), 2,319 (IQR: 1,969-2,780), and 3,582 (IQR: 3,058-4,185) DALYs would be averted with an active microbiology laboratory under the three empirical regimens considered.

### Factors influencing the cost-effectiveness of maintaining an active microbiology laboratory

The key factors that influenced the cost difference and DALYs averted between the intervention and control arm were prevalence of true bacterial infection among suspected bloodstream infections, patient characteristics (i.e. risks of ICU admission), cost of patient care in ICU, cost of maintaining active microbiological testing service, and patient age.

The number of preventable deaths due to an active microbiology laboratory would be higher in a setting with a high prevalence of true bacterial infection, compared to that in a setting with low prevalence (Figure 3). Maintaining an active microbiological testing laboratory service was more cost-saving per unit of increase in health benefit (smaller ICER and higher cost-effectiveness) in a setting with a high prevalence compared to a low prevalence of true bacterial infection. This could be explained by a proportionally increased number of true bacterial infections diagnosed by microbiological testing that would lead to timely appropriate treatments. In other words, the impact of overall delays in appropriate antibiotic treatment would be expected to increase proportionally with the increased number of true bacterial infections. The observation was further supported by the wider gaps in DALYs between the scenarios when narrow spectrum empirical antibiotics were used compared to scenarios using broad spectrum empirical antibiotics (meropenem and vancomycin, which have broader coverage compared to ceftriaxone and gentamicin or ampicillin and gentamicin).

**Figure 3.**
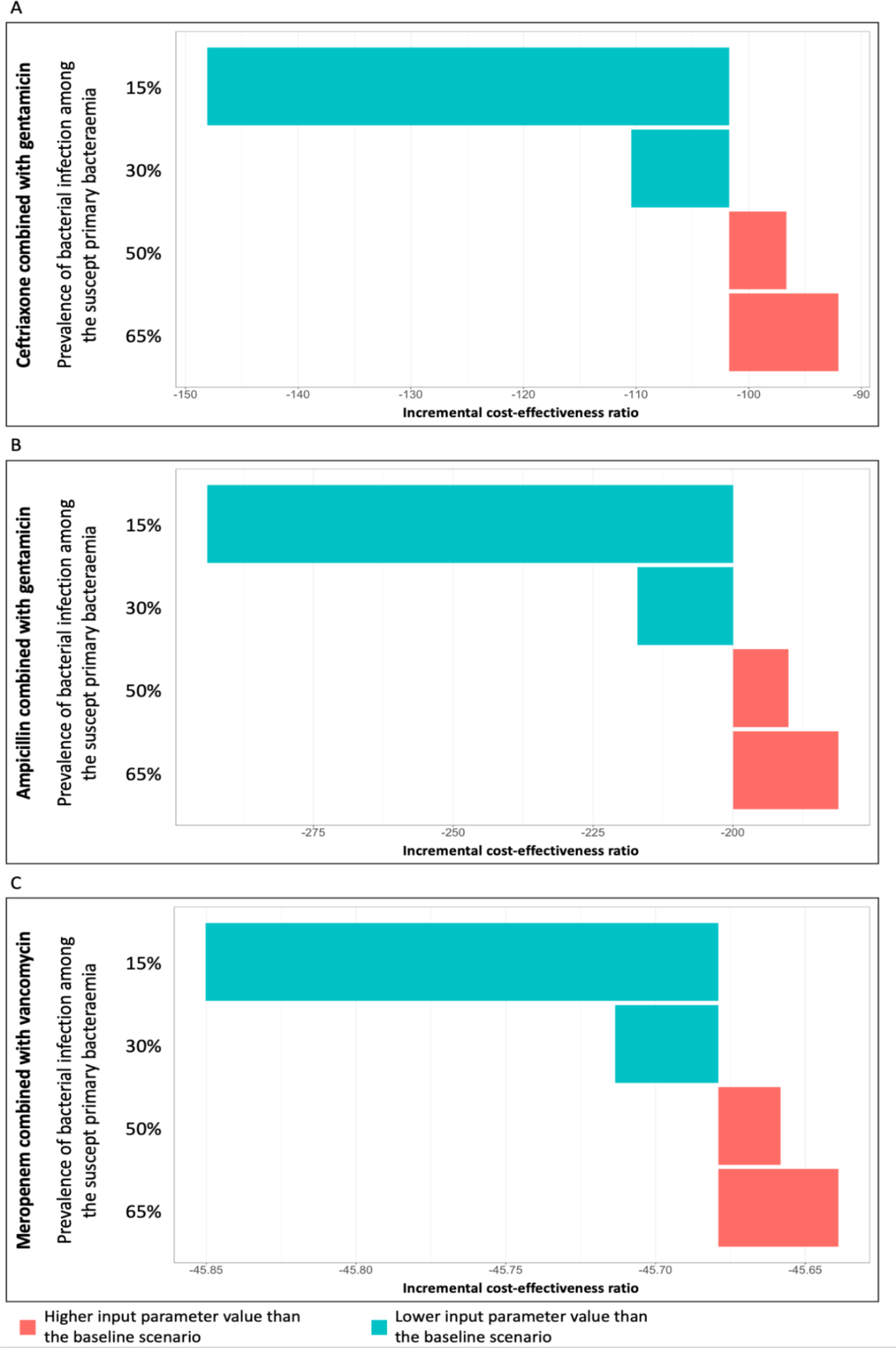
Deterministic sensitivity analysis for parameters related to prevalence of adult bacterial infection under (A) ceftriaxone and gentamicin; (B) ampicillin and gentamicin; and (C) meropenem and vancomycin empirical antibiotic treatment. The baseline scenario assumed that the prevalence of bacterial infection among the suspected primary bacteraemia was 40%. Note the differences in scale of the x-axis in the three plots.

In settings where false negative blood cultures resulted in highly detrimental clinical consequences (equivalent to those experienced by patients with bloodstream infections where the empirical treatment did not cover the causative organisms), the active microbiological testing service arm would still be cost-saving as long as the proportion of false negative is not high (Figure 4). When meropenem and vancomycin were used as empirical antibiotic treatment and probability of culture positive among bacterial infections was as low as 60% and the probability of clinical condition improving among false culture negative (culture results showed negative among patients with bacterial infection) was as low as 1.4%, the ICER would be $57.31, which is less than one GDP per capita ($2,389 the GDP per capita^20^ in 2022 in Timor-Leste), per DALY averted.

**Figure 4.**
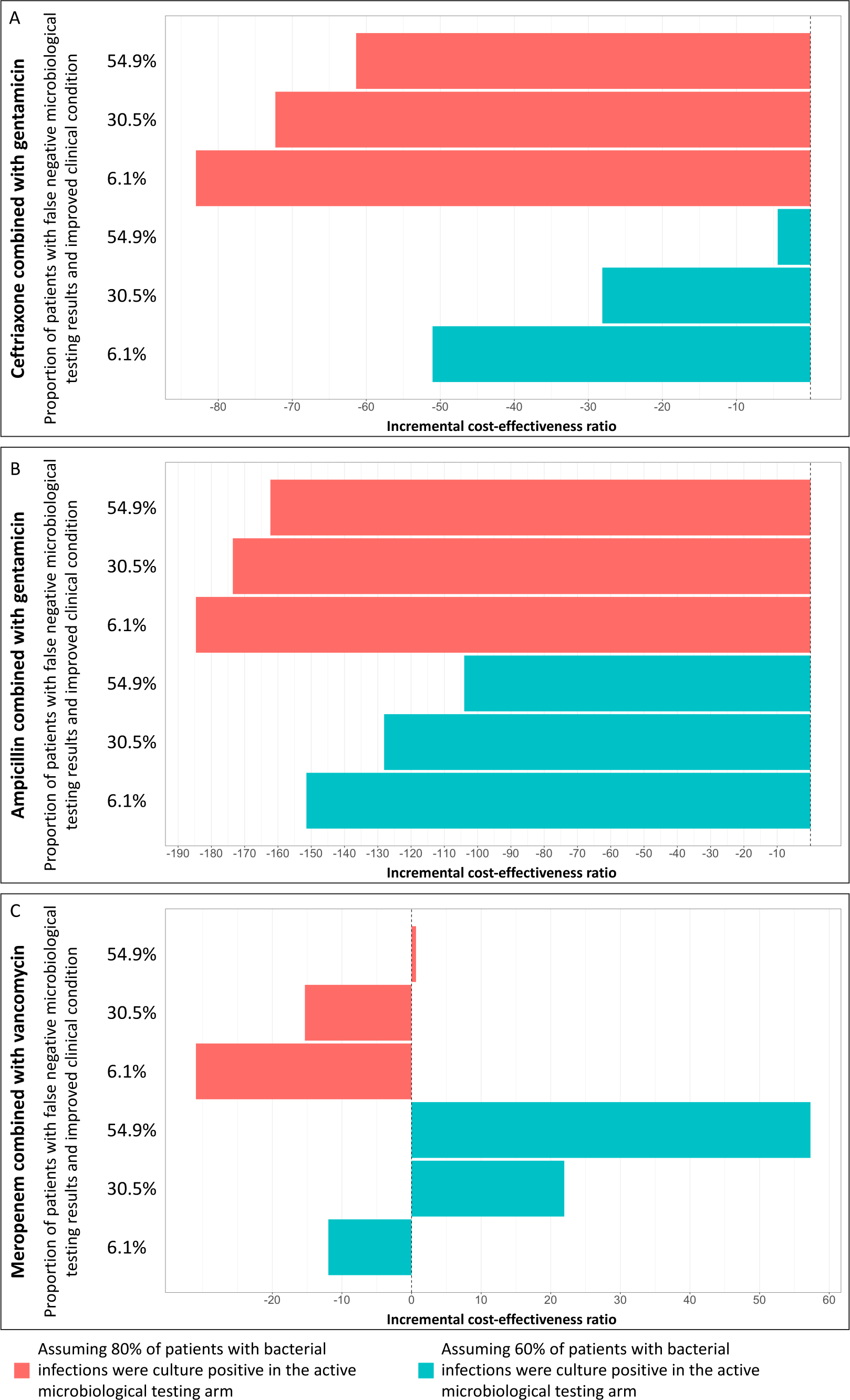
Deterministic sensitivity analyses for parameters on clinical conditions of patients who had false negative cultures under the active microbiological testing arm. The analyses were performed independently for when (A) ceftriaxone and gentamicin; (B) ampicillin and gentamicin; and (C) meropenem and vancomycin were used as empirical antibiotic treatments. The dashed line indicates zero incremental cost-effectiveness. The estimated proportion of patients with suspected primary bacteraemia and false negative blood cultures whose clinical condition deteriorated under the scenarios of ceftriaxone and gentamicin, ampicillin and gentamicin, and meropenem and vancomycin treatment were 39%, 39%, and 26%, respectively, based on the expert elicitations that meropenem and vancomycin treatment would have higher coverage rate than the other two treatment strategies.

When the cost of maintaining the microbiological testing service exceeds $76 per sample, performing microbiological testing on hospitalised patients with suspected primary bloodstream infection with empirical meropenem and vancomycin treatment would cost slightly more than not performing microbiological testing with an estimated ICER of 3.78, which is less than one GDP per capita, per one unit of DALY averted. Similarly, sensitivity analyses assuming two sets of blood samples, which is equivalent to four sample bottles, found that maintaining the microbiological testing service is cost-effective and cost-saving under the scenarios in which ceftriaxone and gentamicin, and ampicillin and gentamicin were used as empirical treatments (Appendix 4). Empirical meropenem and vancomycin treatment would cost $22.33 more per hospitalised adult patient with suspected primary bacteraemia compared to no microbiology testing, with an estimated ICER of $13.31 per DALY averted, which is less than one GDP per capita ($2,389 the GDP per capita^20^ in 2022 in Timor-Leste).

## Discussion

In the current study we modelled the cost-effectiveness of providing a blood culture service for HNGV in Timor-Leste, compared to not having this service. The model outputs in this study, even with conservative parameter values, suggest that maintaining a microbiology laboratory service for this hospital is cost-effective in this lower middle-income country. The microbiology service in this setting may be particularly important in reducing deaths in settings where the prevalence of bacterial infection among suspected bloodstream infection is high.

If the sensitivity of microbiological testing is low (with high rates of false negative tests), the cost effectiveness of maintaining a microbiology laboratory service would be reduced due to a higher proportion of patients needing ICU care due to delayed appropriate treatment. We found that if the sensitivity of microbiological testing was less than 60% and the probability of clinical condition improving in patients who were false culture negative was 1.4%, even under a low cost-effectiveness per DALY averted threshold of 2.5% GDP per capita in Timor-Leste (with $2,389 of the GDP per capita^20^ in 2022) maintaining a microbiology laboratory service would be cost-effective compared to no microbiology testing. The cost-effectiveness thresholds estimated by Woods et al^21^ approach, which adjusted for purchasing power parity, were 1-51% GDP per capita ($1-116) for Malawi, 4-51% GDP per capita ($44-518) for Cambodia, and 14-51% ($472-1,786) for Indonesia.^21^ Accordingly, our results would be likely to viewed as cost-effective in Timor Leste. Steps to ensure high sensitivity of microbiological testing, including training focused on sample collection prior to antibiotics, and collection of adequate blood volumes for blood cultures, as well as laboratory quality assurance factors, remain important to ensure effectiveness treatment and optimise the value of microbiology testing. The US Centers for Disease Control and Prevention (CDC) recommended at least two blood culture sets (four bottles) should be obtained within a few hours of each other via peripheral venipuncture to increase sensitivity in detecting pathogens.^17^ A concern is whether such recommendations, which require additional costs, would still be cost-effective in resource-limited settings. The sensitivity analysis from the current study indicates that while increasing the number of bottles will increase costs substantially, the microbiological testing intervention would still be cost-effective (appendix 4).

Based on the decision tree constructed, a narrower difference in costs between the microbiological testing service and no microbiological testing arms was observed in settings where meropenem and vancomycin were used to treat patients with suspected primary bloodstream infection compared to settings where ceftriaxone and gentamicin, or ampicillin and gentamicin were used. This was due to the high costs of meropenem and vancomycin compared to the other empirical antibiotic regimens considered ($35.16 per day for meropenem and vancomycin versus $1.76 per day for ceftriaxone and gentamicin) and the fact that stepping-up to antibiotics such as colistin (assuming these antibiotics could be available in the near future in Timor-Leste) would impose large treatment costs. Moreover, based on the expert elicitation, empirical treatment with meropenem and vancomycin was less likely to result in deteriorating clinical condition compared with treatment with ceftriaxone and gentamicin or ampicillin and gentamicin. This in turn reduced costs of care and narrowed the cost gap between microbiological testing and no microbiological testing arms in settings where meropenem and vancomycin was appropriately used as empirical antibiotic treatment, noting the risks of contributing to further antimicrobial resistance with inappropriate empiric broad spectrum antibiotic use.

Since 2019, Timor-Leste has participated in the Fleming Fund project, a UK Government funded initiative which supports low- and middle-income countries to generate, share and use data to improve antimicrobial use and encourage investment in the prevention of antimicrobial resistance.^18^ Consequently, Timor-Leste has undergone significant laboratory strengthening activities including improvements to infrastructure, diagnostic test availability, and standardization of microbiology results. These activities combined with increased awareness of the benefits of surveillance have led to a significant increase in utilization of the diagnostic microbiology service. Health economic analysis of the cost effectiveness of a microbiology laboratory service is helpful for informing decision-making regarding ongoing government investments in tackling AMR in Timor-Leste.

There are limitations in this model. Firstly, the clinical inputs are not based on actual, prospectively collected patient data from the national hospital. We have assumed the same mortality rates for patients with bloodstream infections receiving different antibiotic regimens regardless of age (children and adults), and have assumed that having microbiological testing results available will lead to changes in antibiotic prescribing. These assumptions may not always be true, especially if the trust in microbiological testing results among antibiotic prescribers is low and antibiotic prescribing behaviour is influenced by alternate prescriber preferences and beliefs. Secondly, the estimated cost saving in the active microbiological testing arm was driven largely by ICU costs. However, the assumption that reduction in ICU admissions in a given hospital translates proportionally to a reduction in costs when bed capacity and staffing level is maintained may not hold for all settings. Furthermore, the optimal method to cost hospital bed stays is an area of ongoing research.^19^ Thirdly, the long-term population and individual-level impact of antibiotic overuse were not systematically accounted for in this analysis.

## Conclusion

The findings suggest that maintaining a microbiology laboratory service compared to no microbiological testing is likely to be cost-saving in the HNGV tertiary referral hospital in Timor-Leste, both reducing mortality and saving costs. These benefits primarily result from the fact that a microbiological diagnostic service enables severe bacterial infections to be treated with appropriate antibiotics in a timely manner which is expected to result in fewer ICU admissions and lower mortality. These results would be helpful for informing decision-making regarding ongoing government investments in tackling AMR in Timor-Leste.

## Data Availability

All data produced in the present work are contained in the manuscript

## Acknowledgement

We would like to thank all the experts who participated in the expert elicitation exercises.

## Appendices

https://docs.google.com/document/d/1ybpT7FVaVBzQ6O0khMl8_AyJIOdkl5FFpe-nkvLPVMo/edit?usp=sharing

https://docs.google.com/document/d/1NDVcRsrVhhqbz7wWOCQITheJPyXvPALWx_kKxqbIYnY/edit?usp=sharing

